# Childhood Asthma and COVID-19: A Nested Case-Control Study

**DOI:** 10.1101/2021.09.20.21263838

**Authors:** Kristina Gaietto, Megan Culler Freeman, Leigh Anne DiCicco, Sherry Rauenswinter, Joseph R Squire, Zachary Aldewereld, Jennifer Iagnemma, Brian T Campfield, David Wolfson, Traci M Kazmerski, Erick Forno

## Abstract

**Background:** Most pediatric studies of asthma and COVID-19 to date have been ecological, which offer limited insight. We evaluated the association between asthma and COVID-19 at an individual level.

**Methods:** Using data from prospective clinical registries, we conducted a nested case-control study comparing three groups: children with COVID-19 and underlying asthma (“A+C” cases); children with COVID-19 without underlying disease (“C+” controls); and children with asthma without COVID-19 (“A+” controls).

**Results:** The cohort included 142 A+C cases, 1110 C+ controls, and 140 A+ controls. A+C cases were more likely than C+ controls to present with dyspnea and wheezing, to receive pharmacologic treatment including systemic steroids (all p<0.01), and to be hospitalized (4.9% vs 1.7%, p=0.01). In the adjusted analysis, A+C cases were nearly 4 times more likely to be hospitalized than C+ controls (adjusted OR=3.95 [95%CI=1.4-10.9]); however, length of stay and respiratory support level did not differ between groups. Among A+C cases, 8.5% presented with an asthma exacerbation and another 6.3% developed acute exacerbation symptoms shortly after testing positive for SARS-CoV-2. Compared to historic A+ controls, A+C cases had less severe asthma, were less likely to be on controller medications, and had better asthma symptom control (all p<0.01).

**Conclusions:** In our cohort, asthma was a risk factor for hospitalization in children with COVID-19, but not for worse COVID-19 outcomes. SARS-CoV-2 does not seem to be a strong trigger for pediatric asthma exacerbations. Asthma severity was not associated with higher risk of COVID-19.

**Key messages:** In this pediatric cohort, asthma was a risk factor for hospitalization in children with COVID-19, but not for worse COVID-19 outcomes. Baseline asthma severity was not associated with higher risk of COVID-19, and SARS-CoV-2 did not seem to be a strong trigger for pediatric asthma exacerbations.

## INTRODUCTION

There have been conflicting reports on whether asthma increases COVID-19 risk or severity^(1-6)^, with scarce data in children^(7)^. While the majority of COVID-19 cases have occurred in adults^(8)^, pediatric COVID-19 cases in the U.S. have been steadily increasing since July 2021^(9)^. The rise of the highly infectious Delta variant coincides with the return of millions of unimmunized students to in-person classrooms this fall with highly variable mitigation strategies, making it critically important that we better elucidate the impact of COVID-19 on children, particularly those with common chronic diseases such as asthma.

Most pediatric studies on COVID-19 and asthma to date have been epidemiological or ecological, which examine associations at a large scale but provide limited inferences into the individual characteristics driving the findings. Similarly, there have been ecological reports describing reduced pediatric asthma exacerbations and morbidity during the pandemic^(10-13)^, likely as a result of physical distancing, masks, and perhaps other factors such as decreases in air pollution. Yet, those have focused on the concurrent effects of the pandemic on population-level asthma morbidity and healthcare utilization, rather than the direct potential associations between asthma and incident COVID-19 characteristics in children.

In this study, we examined the association between asthma and COVID-19 in children using nested case-control analyses. We hypothesized that (1) there is an association between asthma and COVID-19 presentation and outcomes, and (2) that asthma severity is correlated with COVID-19 risk.

## METHODS

### Study population and data collection

The Western Pennsylvania COVID-19 Registry (WPACR) is a secure database established in March 2020 to record baseline characteristics, acute presentation, and initial outcomes of pediatric patients (ages 0-21 years) presenting with a SARS-CoV-2 infection to UPMC Children’s Hospital of Pittsburgh (CHP, the largest pediatric referral center in the region) and Children’s Community Pediatrics (CCP, the associated primary care network)^(14)^. For this analysis, we extracted data from children with pre-existing asthma who presented with COVID-19 between March and December 2020 (“A+C” cases). As disease controls, we selected from the WPACR children without pre-existing conditions who presented with COVID-19 (“C+” controls) during the same period; as well as children with asthma (“A+” controls) recruited to the CHP Asthma Registry during the same period the year prior to the pandemic (March to December 2019). The Asthma Registry includes children seen for asthma in the CHP Pulmonary or Allergy clinics, the Emergency Department, who are hospitalized for asthma, or who participate in asthma research studies at our center.

We have previously described details of the WPACR^(14)^; in brief, we included subjects if they had a positive SARS-CoV-2 RT-PCR or if they met criteria for the multisystem inflammatory syndrome in children (MIS-C), and a multidisciplinary team representing the different pediatric specialties involved in the care of patients with COVID-19 at our institution (including primary care, pulmonology, hospital medicine, adolescent medicine, infectious disease, and critical care medicine) extracted relevant clinical data from the EHR. Data abstracted included patient demographics, symptoms and initial presentation, healthcare utilization data, laboratory results, and acute disease outcomes.

For all patients with asthma (A+C cases and A+ controls), we also directly abstracted EHR data on baseline asthma severity, asthma controller medications, symptom control (Asthma Control Test [ACT]^(15)^ scores for adolescents ≥12 years old or Childhood Asthma Control Test [C-ACT]^(16)^ scores for children 4-11 years old), lung function (FEV1 and FVC as percent-of predicted [%pred], as well as FEV1/FVC ratio), and atopy biomarkers (total and specific IgE, allergy skin testing, and peripheral blood eosinophil counts). We defined asthma severity based on National Asthma Education and Prevention Program (NAEPP) guidelines^(17)^; and poor control as an ACT or C-ACT ≤19. We evaluated asthma exacerbations in both the A+C and A+ groups from January 1, 2018, to December 31, 2019, to avoid any potential impact of the pandemic on asthma control, management, or health care utilization.

The WPACR and the CHP Asthma Registry are both approved by the Institutional Review Board at the University of Pittsburgh (protocols STUDY20110072 and STUDY19020359, respectively).

### Statistical Analysis

For the analysis of A+C cases vs C+ controls (i.e., the differences in COVID-19 between children with and without asthma), we compared initial symptoms at presentation, history of recent travel, and known exposures. Our primary outcomes of interest were hospitalization, hospital length-of-stay (LOS), PICU admission, and the maximal respiratory support required during hospitalization. For the analysis between A+C cases and A+ controls (i.e., to assess whether patients presenting with COVID-19 had more severe pre-existing asthma than expected from our usual hospital population), our primary variables were asthma severity and symptom control; secondary characteristics were asthma exacerbations, lung function, and atopy biomarkers. We conducted bivariate analyses using 2-tailed t tests or Wilcoxon rank-sum (Mann-Whitney U) tests for continuous variables, and chi-squared or Fisher’s exact tests for categorical variables. We conducted adjusted analyses using logistic regression for categorical variables (e.g., hospitalization), Poisson regression for count data (e.g., number of severe asthma exacerbations), or linear regression for continuous variables. We adjusted models for hospitalization for age, sex, and covariates that were significant in the bivariate analyses of asthma and hospitalization: race, BMI, days from symptom onset to presentation, and non-asthma symptoms (fever, fatigue, and vomiting). We performed all analyses using STATA v16.1 (StataCorp, College Station, TX) or SAS v9.4 (SAS Institute, Cary, NC).

## RESULTS

From March 11, 2020, to December 21, 2020, there were 1,802 cases with SARS-CoV-2 RT-PCR+ in the WPACR (**Figure 1**). Of those, 142 had asthma and were extracted as A+C cases; and 1,110 had no reported pre-existing conditions and were selected as C+ controls. We also extracted 140 asthma controls (A+ controls) recruited to the CHP Asthma Registry during the same period in 2019.

**Figure 1.**
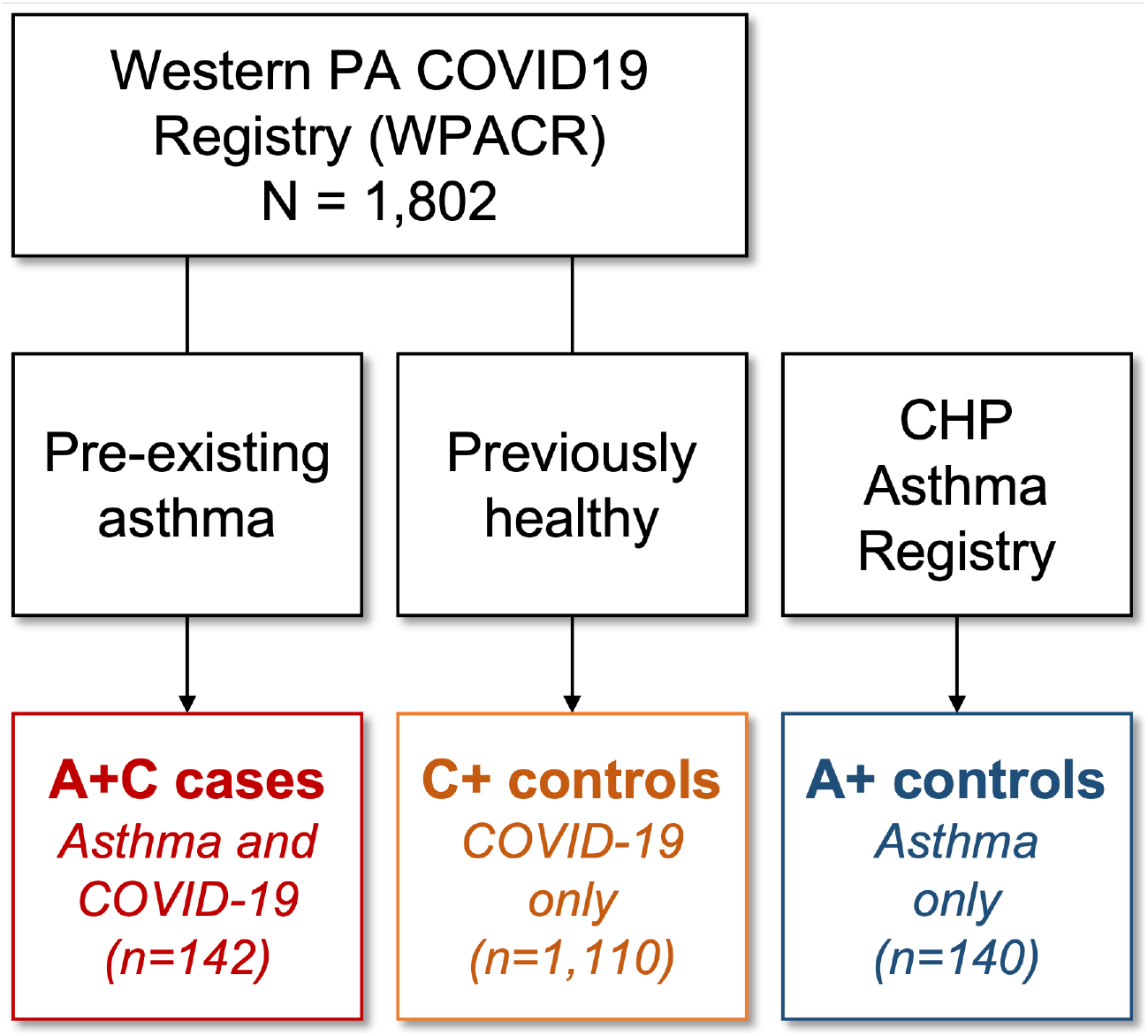
Diagram of study cohort selection. A+C cases and C+ controls were recruited from March to December 2020. A+ controls were selected from patients recruited from March to December 2019. Data on asthma characteristics (for both A+C cases and A+ controls) was collected for 2018-2019 to avoid any potential impact of the pandemic on asthma control, management, or health care utilization.

Baseline cohort characteristics are shown in **Table 1**. A+C cases (COVID-19 with pre-existing asthma) were approximately 4 years older than both C+ and A+ controls (median ages 14.6, 12.0, and 10.2 years, respectively), but there were no differences in sex distributions between the three groups. Compared to children without asthma, those with asthma (with or without COVID-19) were more likely to be Black (A+C cases 25.4% and A+ controls 23.6%, vs C+ controls 10.2%) and had a higher BMI (median BMI percentiles 79.0 and 78.6, respectively, vs 63.0).

**Table 1.** Demographic characteristics of the participants in the study.

|  | **A+C Cases**<br>(Asthma and COVID-19) | **C+ Controls**<br>(COVID-19 only) | **P-value** | **A+ Controls**<br>(Asthma only) | **P-value** |
| --- | --- | --- | --- | --- | --- |
| N | 142 | 1,110 |  | 140 |  |
| Age, years | 14.6 [10.7-17.9] | 12.0 [4.8-16.9] | **<0.001** | 10.2 [7.8-13.9] | **<0.001** |
| Male sex | 79 (55.6%) | 560 (50.5%) | 0.24 | 87 (62.1%) | 0.27 |
| Race: |  |  | **<0.001** |  | 0.88 |
| - White | 103 (72.5%) | 905 (81.5%) |  | 103 (73.6%) |  |
| - Black | 36 (25.4%) | 113 (10.2%) |  | 33 (23.6%) |  |
| - Other/unknown | 3 (2.1%) | 92 (8.3%) |  | 4 (2.9%) |  |
| Hispanic | 1 (0.7%) | 8 (0.7%) | 0.96 | 7 (5%) | **0.03** |
| BMI, percentile | 79.0 [50.0-95.2] | 63.0 [30.6-87.6] | **<0.001** | 78.6 [48.1-95.3] | 0.93 |

Numbers represent mean (SD) or median [interquartile range] for continuous variables, and n (%) for categorical variables. BMI: Body mass index. LOS: Length of stay (days). P-values for the comparison of each control group vs A+C cases.

### COVID-19 hospitalizations

Altogether, 26 children (2.1%) in the WPCAR were hospitalized during the study period (**Supplementary Table S1**), including 9 PICU admissions and no deaths. Compared to children with COVID-19 who were not hospitalized, hospitalized patients were more likely to have asthma (26.9% vs 11.0%). They were also younger (median ages 7.4 vs 12.6 years); more likely to be Black (38.5% vs 12.3%); to present with fever, fatigue, wheezing, dyspnea, chest pain, and/or vomiting; and to receive pharmacologic treatment (38.5% vs 1.2%) (**Supplementary Table S1**). PICU admissions were few but included 1/7 A+C cases and 8/19 C+ controls.

### COVID-19 in children with asthma vs. no underlying disease

A+C cases were more likely than C+ controls to endorse recent travel; to present with wheezing, dyspnea, chest pain, and loss of taste; and to be hospitalized (4.9% vs 1.7%) (**Table 2**). They were also more likely to receive pharmacologic treatment including albuterol and systemic steroids. Among the 142 A+C cases, 12 (8.5%) had an asthma exacerbation as part of the initial presentation that led to SARS-CoV-2 testing, and an additional 9 (6.3%) developed an asthma exacerbation shortly after they tested positive for COVID-19. In the remaining 118 cases, asthma was either solely a pre-existing condition with no active symptoms upon COVID-19 presentation (80.1%) or there was insufficient data to determine asthma symptoms (4.2%). A+C cases presenting with acute asthma symptoms were more likely to be hospitalized than those with no acute asthma symptoms (3/21 [14.3%] vs 3/118 [2.5%], p=0.04).

**Table 2.** COVID-19 characteristics in COVID-19 cases (A+C) and (C+) controls the study.

|  | **A+C Cases** | **C+ Controls** | **P-value** |
| --- | --- | --- | --- |
| Initial presentation: |  |  | 0.58 |
| - ED or urgent care | 19 (13.4%) | 113 (10.2%) |  |
| - Primary care or telemedicine | 122 (85.9%) | 974 (87.8%) |  |
| - Other/unknown | 1 (0.7%) | 23 (2.1%) |  |
| Recent travel | **10 (7.0%)** | **34 (3.1%)** | **0.009** |
| Known exposure | 86 (62.3%) | 746 (68.3%) | 0.16 |
| Interval, days: |  |  |  |
| - Symptoms to presentation | 2 [1-3] | 2 [1-3] | 0.37 |
| - Symptoms to test | 2.5 [1-4] | 2 [1-4] | 0.47 |
| Initial symptoms: |  |  |  |
| - Asymptomatic | 16 (11.3%) | 124 (11.2%) | 0.97 |
| - Fever | 53 (37.3%) | 443 (39.9%) | 0.55 |
| - Fatigue | 24 (16.9%) | 162 (14.6%) | 0.47 |
| - Cough | 62 (43.7%) | 477 (43.0%) | 0.88 |
| - Wheezing | **7 (4.9%)** | **5 (0.5%)** | **<0.001** |
| - Dyspnea | **13 (9.2%)** | **25 (2.3%)** | **<0.001** |
| - Chest pain | **7 (4.9%)** | **16 (1.4%)** | **0.004** |
| - Loss of smell | 23 (16.2%) | 125 (11.3%) | 0.09 |
| - Loss of taste | **26 (18.3%)** | **129 (11.6%)** | **0.02** |
| - Abdominal pain | 8 (5.6%) | 52 (4.7%) | 0.62 |
| - Vomiting | 6 (4.2%) | 36 (3.2%) | 0.54 |
| Initial treatment: |  |  |  |
| - Any pharmacologic treatment | **10 (7%)** | **15 (1.3%)** | **<0.001** |
| - Albuterol | **10 (7%)** | **8 (0.7%)** | **<0.001** |
| - Systemic steroids | **5 (3.5%)** | **10 (0.8%)** | **0.007** |
| **Main outcomes** |  |  |  |
| Hospitalized | **7 (4.9%)** | **19 (1.7%)** | **0.01** |
| Hospital LOS (days) | 1 [1-4] | 2 [1-4] | 0.51 |
| Respiratory support<sup>†</sup>: |  |  | 0.72 |
| - None (room air) | 6 | 14 |  |
| - Nasal cannula (NC) | 0 | 2 |  |
| - High-flow NC or NPAP | 1 | 2 |  |
| - Invasive ventilation | 0 | 1 |  |

Numbers represent mean (SD) or median [interquartile range] for continuous variables, and n (%) for categorical variables.
<sup>†</sup>Among hospitalized patients. LOS: Length of stay (days).

In the multivariable analysis (**Table 3**), asthma was a risk factor for hospitalization with COVID-19: after adjustment for age, sex, race, recent travel and known exposure, A+C cases were nearly 4 times more likely to be hospitalized than C+ controls (aOR=3.95 [95%CI=1.4-10.9]; **Table 3 Model 1**). Results were similar after additionally adjusting for BMI percentile and the number of days between symptom onset and presentation (**Table 3 Model 2**), or after adjusting for other, non-asthma symptoms that were associated with hospitalization (fever, fatigue, and vomiting; **Table 3 Model 3**). Among hospitalized patients, however, the length of stay and respiratory support level did not significantly differ by asthma status (**Table 2**).

**Table 3.** Association between asthma and hospitalization for COVID-19.

|  | **Model 1**<br>(N=1,252) | **Model 2**<br>(N=873) | **Model 3**<br>(N=1,252) |
| --- | --- | --- | --- |
| **Asthma** | **3.95 (1.43-10.9)\*\*** | **4.43 (1.35-14.6)\*\*** | **3.33 (1.19-9.33)\*** |
| Age, years | **0.88 (0.82-0.95)\*\*** | **0.90 (0.82-0.99)\*** | **0.91 (0.85-0.98)\*** |
| Male sex | 0.55 (0.24-1.26) | 0.39 (0.13-1.17) | 0.49 (0.21-1.15) |
| Race: |  |  |  |
| - White | Ref. | Ref. | Ref. |
| - Black | **3.00 (1.28-7.07)\*** | 2.53 (0.74-8.67) | **4.55 (1.87-11.1)\*** |
| - Other/unknown | 0.54 (0.07-4.23) | 2.04 (0.24-17.2) | 0.51 (0.06-4.18) |
| Recent travel | 1.39 (0.52-3.68) | 2.42 (0.26-22.6) | 1.34 (0.40-4.52) |
| Known exposure | **0.27 (0.52-0.62)\*\*** |  |  |
| BMI, percentile |  | 1.01 (0.99-1.03) |  |
| Days from symptom onset to presentation |  | 1.002 (0.99-1.01) |  |
| Initial symptoms: |  |  |  |
| - Fever |  |  | 1.90 (0.40-4.52) |
| - Fatigue |  |  | **3.15 (1.23-8.09)\*** |
| - Vomiting |  |  | **8.62 (3.15-23.6)\*\*** |

### Comparison between cases and asthma controls

In our evaluation of whether children with asthma presenting with COVID-19 differ from children with asthma usually seen at our center (**Table 4**), A+C cases had less severe asthma (58% vs 23% had intermittent asthma) and lower eosinophil counts (median 110 vs 300 cells/μL), and they were less likely to be on controller medications, and less likely to have poor symptom control (13.6% vs 30.7%). This was driven by adolescent A+C cases who had better ACT scores compared to A+ controls (mean 22.1 vs 20.3), with no differences in C-ACT scores among the younger children. There were no differences in lung function (FEV1, FVC, or FEV1/FVC), but A+C cases were less likely to have atopic comorbidities, and they had lower rates of severe exacerbations compared to A+ controls (median 1.0 vs 1.5 events per year, Mann Whitney p=0.042); this difference persisted after adjustment for age, sex, and race (difference -0.43 events per year [95%CI -0.77 to -0.10], Poisson p=0.01).

**Table 4.**
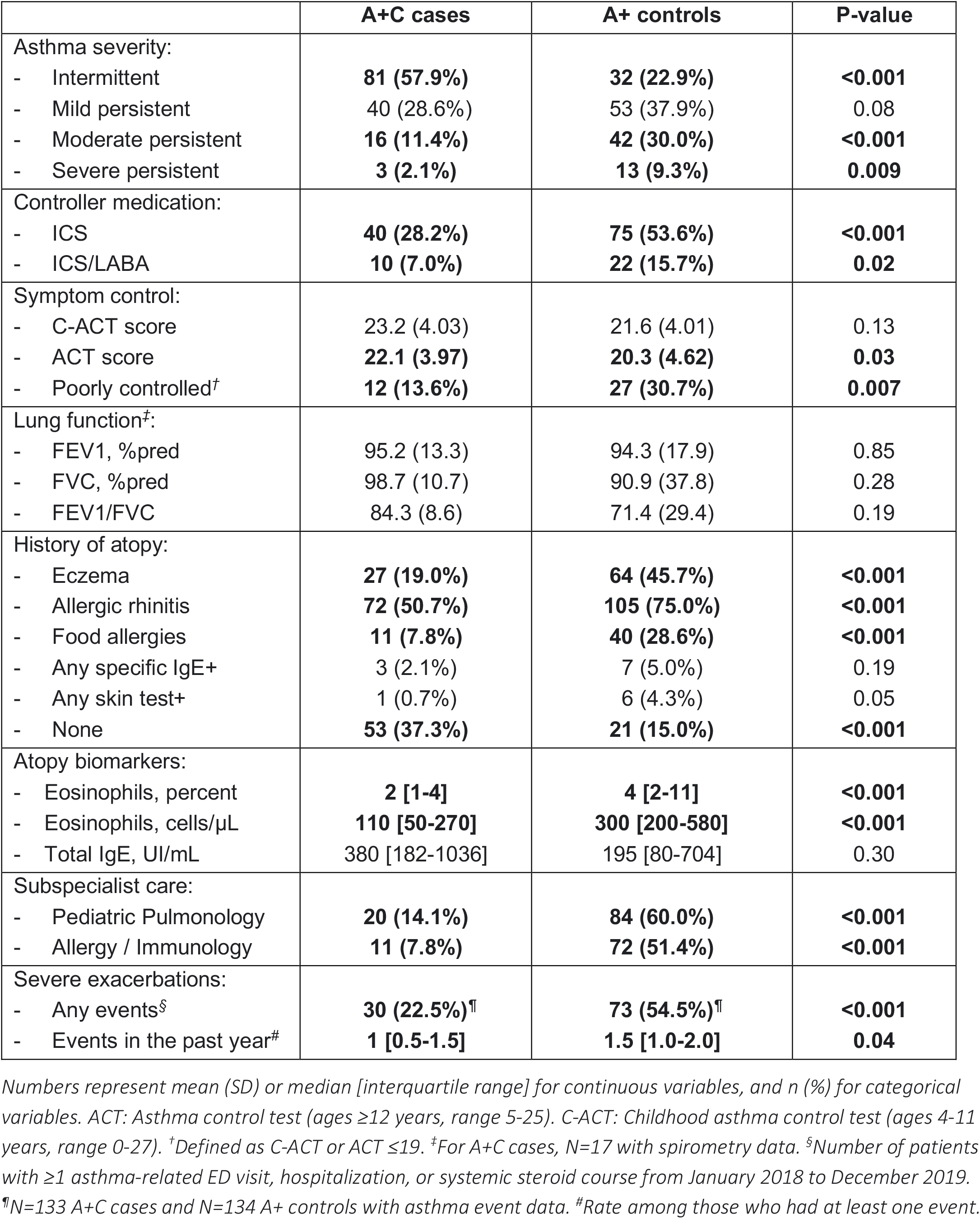
Asthma characteristics in COVID-19 cases (A+C) and asthma (A+) controls.

|  | A+C cases | A+ controls | P-value |
| --- | --- | --- | --- |
| Asthma severity: |  |  |  |
| - Intermittent | **81 (57.9%)** | **32 (22.9%)** | **<0.001** |
| - Mild persistent | 40 (28.6%) | 53 (37.9%) | 0.08 |
| - Moderate persistent | **16 (11.4%)** | **42 (30.0%)** | **<0.001** |
| - Severe persistent | **3 (2.1%)** | **13 (9.3%)** | **0.009** |
| Controller medication: |  |  |  |
| - ICS | **40 (28.2%)** | **75 (53.6%)** | **<0.001** |
| - ICS/LABA | **10 (7.0%)** | **22 (15.7%)** | **0.02** |
| Symptom control: |  |  |  |
| - C-ACT score | 23.2 (4.03) | 21.6 (4.01) | 0.13 |
| - ACT score | **22.1 (3.97)** | **20.3 (4.62)** | **0.03** |
| - Poorly controlled<sup>†</sup> | **12 (13.6%)** | **27 (30.7%)** | **0.007** |
| Lung function<sup>‡</sup>: |  |  |  |
| - FEV1, %pred | 95.2 (13.3) | 94.3 (17.9) | 0.85 |
| - FVC, %pred | 98.7 (10.7) | 90.9 (37.8) | 0.28 |
| - FEV1/FVC | 84.3 (8.6) | 71.4 (29.4) | 0.19 |
| History of atopy: |  |  |  |
| - Eczema | **27 (19.0%)** | **64 (45.7%)** | **<0.001** |
| - Allergic rhinitis | **72 (50.7%)** | **105 (75.0%)** | **<0.001** |
| - Food allergies | **11 (7.8%)** | **40 (28.6%)** | **<0.001** |
| - Any specific IgE+ | 3 (2.1%) | 7 (5.0%) | 0.19 |
| - Any skin test+ | 1 (0.7%) | 6 (4.3%) | 0.05 |
| - None | **53 (37.3%)** | **21 (15.0%)** | **<0.001** |
| Atopy biomarkers: |  |  |  |
| - Eosinophils, percent | **2 [1-4]** | **4 [2-11]** | **<0.001** |
| - Eosinophils, cells/μL | **110 [50-270]** | **300 [200-580]** | **<0.001** |
| - Total IgE, UI/mL | 380 [182-1036] | 195 [80-704] | 0.30 |
| Subspecialist care: |  |  |  |
| - Pediatric Pulmonology | **20 (14.1%)** | **84 (60.0%)** | **<0.001** |
| - Allergy / Immunology | **11 (7.8%)** | **72 (51.4%)** | **<0.001** |
| Severe exacerbations: |  |  |  |
| - Any events<sup>§</sup> | **30 (22.5%)<sup>¶</sup>** | **73 (54.5%)<sup>¶</sup>** | **<0.001** |
| - Events in the past year<sup>#</sup> | **1 [0.5-1.5]** | **1.5 [1.0-2.0]** | **0.04** |

Numbers represent mean (SD) or median [interquartile range] for continuous variables, and n (%) for categorical variables. ACT: Asthma control test (ages $\geq 12$ years, range 5-25). C-ACT: Childhood asthma control test (ages 4-11 years, range 0-27). <sup>†</sup>Defined as C-ACT or ACT $\leq 19$ . <sup>‡</sup>For A+C cases, N=17 with spirometry data. <sup>§</sup>Number of patients with $\geq 1$ asthma-related ED visit, hospitalization, or systemic steroid course from January 2018 to December 2019.
<sup>¶</sup>N=133 A+C cases and N=134 A+ controls with asthma event data. <sup>#</sup>Rate among those who had at least one event.

We then evaluated the subgroup of cases and controls who had been previously seen at one of the asthma subspecialty clinics (17 A+C cases and 86 A+ controls seen in either Pulmonology or Allergy clinics; **Supplementary Table S2**). In the analysis restricted to this subgroup, A+C cases and A+ controls were similar in most asthma characteristics, except for persistent differences in eosinophil counts and the proportion of patients with no atopic comorbidities.

## DISCUSSION

Our study is one of the first individual-level, case-control studies on pediatric COVID-19 and asthma. We evaluated the association between asthma and COVID-19 presentation and outcomes in children, and whether asthma severity is correlated with COVID-19 risk, by comparing cases with asthma and acute COVID-19 to controls with COVID-19 only (without asthma) and asthma only (without COVID-19). To our knowledge, it is also one of the first studies to report the incidence of asthma exacerbations among children with asthma with acute SARS-CoV-2 infection.

Of the 1,802 patients in our pediatric COVID-19 registry during the study period, 142 (7.9%) had asthma. In our initial report over the first five months of the pandemic, asthma cases comprised 10.6% of the registry^(14)^. Thus, asthma does not appear to be over-represented among pediatric cases of COVID-19 compared to population asthma prevalence at the national (7.0%)^(18)^ or local (8.0%)^(19)^ levels. Consistent with our results, a retrospective study in Philadelphia revealed an asthma prevalence among pediatric COVID-19 cases there comparable to the local pediatric asthma prevalence estimates^(20)^. A recent retrospective study from Turkey reported that, among children with COVID-19 at their institution, only 54 (<1%) had asthma^(21)^. When compared to 162 controls (COVID-19 without asthma), asthma was associated with higher risk of hospitalization; however, there were no adjusted estimates after accounting for potential confounders, and the low prevalence raises questions about the generalizability of these findings to children in other countries. Other studies have reported asthma prevalence rates among pediatric COVID-19 cases to vary from 0.5% (Wuhan, China)^(22)^ to 20% (Washington, DC)^(23)^, and from 2% (Lombari and Luguria, Italy)^(24)^ to 24% (New York City)^(25)^ among admitted pediatric COVID-19 patients. This may be partly explained by variability in the local asthma prevalence, though at least one study in Brazil found a lower asthma prevalence rate among children with COVID-19 infection compared to the local general pediatric asthma prevalence (13% vs 20-25%), suggesting that there may be other contributors to the variability among studies^(26)^.

In our cohort, children with co-morbid asthma were nearly 4 times more likely to be hospitalized for COVID-19 compared to children without asthma, even after adjusting for relevant covariates and potential confounders like age, sex, race/ethnicity, BMI, recent travel, known exposures, and the number of days between symptom development and the patient’s initial presentation to care. Results remained essentially unchanged after further adjusting for non-asthma related symptoms that could have independently increased likelihood of hospitalization, such as fever, vomiting, and fatigue. Consistent with our results, a previous study in Colorado reported asthma as a risk of hospitalization in children with COVID-19, along with other chronic conditions^(27)^. However, others have reported conflicting results^(20, 23)^. Given the paucity of individual-level studies evaluating the risk of asthma in COVID-19 among children, certain providers or systems may have had differing thresholds to admit a child with asthma compared to one without pre-existing conditions. Once hospitalized, however, the length of hospital stay and the need for respiratory support did not significantly differ by asthma diagnosis. Similarly, while we had too few PICU admissions to analyze, numbers did not widely differ by asthma status. This further suggests there may have been a different threshold to admit children with asthma.

We specifically evaluated whether children with asthma and COVID-19 presented with symptoms of acute asthma exacerbations, and found that SARS-CoV-2 does not seem to be a particularly strong trigger. Over 80% of children with asthma who had COVID-19 did not have significant asthma symptoms during their acute viral illness. We did find, however, that children presenting with acute asthma symptoms were more likely to be hospitalized than those with no acute symptoms, and that children with asthma were more likely to receive systemic steroids as part of their COVID-19 management. A recent study of adolescents and adults with COVID-19 similarly reported that only 13% of patients with asthma had wheezing, and only 1/77 was admitted for an asthma exacerbation^(28)^. While we cannot rule out that some patients with asthma may have had exacerbations triggered by mild COVID-19 that were managed at home and who never sought medical attention, this is unlikely given that during the study period the majority of individuals with viral and/or respiratory symptoms were likely tested for COVID-19, making this a reasonable estimate of the percentage of SARS-CoV-2 infections among children with asthma that trigger acute exacerbations.

In our analysis of cases vs historic asthma controls, we found that asthma severity was not associated with higher risk of SARS-CoV-2 infection. Due to the nature of our registries, we were unable to compare these patients to children with asthma managed by primary care providers in the community (i.e., without subspecialty referral or recent admissions to our hospital). However, our findings suggest that children with asthma and COVID-19 were at least not as severe as those who would have been usually referred to subspecialty asthma care at our center. When we restricted our analyses to those children with asthma who had been followed by a specialty clinic in the years prior to the pandemic, there was no significant difference in ACT scores or overall asthma control compared to asthma controls. While the sample size for this subgroup analysis was small, it again suggests that asthma severity was no different in children with COVID-19 than what we would have expected from the population our center usually serves.

Obesity has been identified as a risk factor for COVID-19 infection and worse outcomes, particularly in adults^(29)^. Children with asthma had higher BMI in our cohort, but the association between asthma and hospitalization remained significant after adjustment for BMI. Numbers were too small to examine whether overweight/obesity was associated with hospitalization among children with asthma. While not the main focus of our study, it is important to point out that, in our cohort, African American children were more likely to be hospitalized for COVID-19, consistent with multiple prior studies identifying racial disparities among those with more severe COVID-19^(30-32)^. In our cohort, this association remained significant after adjustment for asthma status and other demographic characteristics.

There are several key strengths to this study. It is one of the first studies to compare COVID-19 presentation and outcomes between children with and without asthma using individual-level characteristics rather than ecological data, and is likely the first such study in a cohort from the U.S. There has been a significant knowledge gap regarding COVID-19 outcomes of children with underlying pulmonary diseases; this study starts to fill some of these gaps, which is of utmost importance for counseling families of children with asthma. Data for this study is based on registries in which information is manually abstracted from the EHR by a multidisciplinary team who is involved in the care of patients with COVID-19, providing more detailed, complete, and accurate information than reports based on diagnostic or billing codes. Furthermore, all asthma information for both cases and historic controls was abstracted by a single asthma provider, eliminating concerns for interrater bias. Finally, given our sample size, we were able to evaluate subgroup characteristics, such as those patients with asthma referred to subspecialty care at our center.

Our study also has several limitations. Given the nature of the pandemic, follow-up duration has been short, precluding the ability to track long-term outcomes. However, it seems unlikely that asthma would have a lasting impact on COVID-19 outcomes, given the generally mild courses of the children in this study. Yet, future studies should also focus on long-term consequences such as changes in lung function or asthma severity and control in children who recovered from acute COVID-19. Another limitation is the small number of hospitalized patients in this study; it is unclear if this is secondary to regional variation in severity or management, possibly due to local variants, or if the low proportion of hospitalized children is secondary to our ability to include even mild and asymptomatic cases (e.g. asymptomatic children who tested positive for SARS-CoV-2) in the registry. It will be crucial to analyze multi-center data to evaluate whether findings vary across different settings. More importantly, the current study period preceded the emergence of the Delta variant of the virus; our registry is ongoing and we plan to analyze any potential differences as we see the already surging wave of pediatric cases across the country.

In summary, we found that asthma severity does not seem to be associated with increased risk of SARS-CoV-2 infection in children. Pre-existing asthma did increase the risk of hospitalization for COVID-19 in our population, but hospital length of stay, need for respiratory support, and disposition did not differ from children without asthma. In this cohort, SARS-CoV-2 infection was not commonly a trigger for asthma exacerbations, but children presenting with symptoms of an asthma exacerbation were more likely to be admitted. With the advent of the Delta variant and current rise in COVID-19 cases, it will be important to conduct further, multi-center, individual-level, case-control or cohort studies of COVID-19 and asthma to better understand this evolving disease and its impact on children with asthma.

## Data Availability

Data can be requested from the authors, after final peer-reviewed publication and contingent on the necessary ethics / institutional review board approvals.

---

**Supplementary Table S1.** Characteristics associated with hospitalization.

|  | **Hospitalized**<br>**(N=26)** | **Not hospitalized**<br>**(N=1,226)** | **P-value** |
| --- | --- | --- | --- |
| Age, years | **7.4 [0.3-15.1]** | **12.6 [6.0-17.1]** | **0.004** |
| Male sex | 10 (38.5%) | 629 (51.3%) | 0.19 |
| Race: |  |  | **0.001** |
| - White | **15 (57.7%)** | **993 (81.0%)** |  |
| - Black | **10 (38.5%)** | **139 (11.3%)** |  |
| - Other/unknown | **1 (3.9%)** | **94 (7.7%)** |  |
| Hispanic | 0 (0%) | 9 (0.7%) | 0.59 |
| BMI, percentile (n=960) | 82.0 [33.0-98.5] | 65.8 [32.8-88.8] | 0.08 |
| Pre-existing asthma diagnosis | **7 (26.9%)** | **135 (11.0%)** | **0.01** |
| Recent travel | 1 (3.9%) | 43 (3.5%) | 0.94 |
| Known exposure | **10 (38.5%)** | **822 (67.1%)** | **0.002** |
| Interval, days: |  |  |  |
| - Symptoms to presentation | **3 [1-4]** | **2 [1-3]** | **0.01** |
| - Symptoms to test | 3 [1-5] | 2 [1-4] | 0.71 |
| Initial symptoms: |  |  |  |
| - Asymptomatic | 3 (11.5%) | 137 (11.2%) | 0.95 |
| - Fever | **17 (65.4%)** | **479 (39.1%)** | **0.007** |
| - Fatigue | **8 (30.8%)** | **178 (14.5%)** | **0.02** |
| - Cough | 15 (57.7%) | 524 (42.7%) | 0.13 |
| - Wheezing | **5 (19.2%)** | **7 (0.6%)** | **<0.001** |
| - Dyspnea | **7 (26.9%)** | **31 (2.5%)** | **<0.001** |
| - Chest pain | **3 (11.5%)** | **20 (1.6%)** | **<0.001** |
| - Loss of smell | 6 (23.1%) | 142 (11.6%) | 0.07 |
| - Loss of taste | 5 (19.2%) | 150 (12.2%) | 0.28 |
| - Abdominal pain | 2 (7.7%) | 58 (4.7%) | 0.48 |
| - Vomiting | **7 (26.9%)** | **35 (2.9%)** | **<0.001** |
| Initial treatment: |  |  |  |
| - Any pharmacologic treatment | **10 (38.5%)** | **15 (1.2%)** | **<0.001** |
| - Albuterol | **4 (15.4%)** | **14 (1.1%)** | **<0.001** |
| - Systemic steroids | **7 (26.9%)** | **4 (0.33%)** | **<0.001** |

Numbers represent mean (SD) or median [interquartile range] for continuous variables, and n (%) for categorical variables. BMI: Body mass index.

**Supplementary Table S2.** Asthma characteristics restricted to cases and controls seen in CHP asthma subspecialty clinics (Pulmonology and/or Allergy)

|  | A+C cases | A+ controls | P-value |
| --- | --- | --- | --- |
| Asthma severity: |  |  | 0.70 |
| - Intermittent | 8 (27.6%) | 32 (23.5%) |  |
| - Mild persistent | 13 (44.8%) | 50 (36.8%) |  |
| - Moderate persistent | 6 (20.6%) | 41 (30.2%) |  |
| - Severe persistent | 2 (6.9%) | 13 (9.6%) |  |
| Controller medication: |  |  |  |
| - ICS | 16 (55.2%) | 71 (52.2%) | 0.77 |
| - ICS/LABA | 5 (17.2%) | 22 (16.2%) | 0.89 |
| Symptom control: |  |  |  |
| - C-ACT score | 23.7 (1.5) | 21.6 (4.0) | 0.38 |
| - ACT score | 20.9 (3.6) | 20.2 (4.7) | 0.60 |
| - Poorly controlled<sup>1</sup> | 4 (23.5%) | 27 (31.4%) | 0.77 |
| Lung function: |  |  |  |
| - FEV1, %pred | 95.2 (13.3) | 94.1 (17.7) | 0.80 |
| - FVC, %pred | 98.7 (10.7) | 92.2 (38.2) | 0.46 |
| - FEV1/FVC | 84.3 (8.6) | 72.3 (28.1) | 0.08 |
| History of atopy: |  |  |  |
| - Eczema | 8 (27.6%) | 61 (44.9%) | 0.09 |
| - Allergic rhinitis | 18 (62.1%) | 102 (75.0%) | 0.16 |
| - Food allergies | 6 (20.7%) | 40 (29.4%) | 0.34 |
| - Any specific IgE+ | 3 (10.3%) | 7 (5.2%) | 0.38 |
| - Any skin test+ | 1 (3.5%) | 6 (4.4%) | 0.99 |
| - None | **9 (31.0%)** | **21 (15.4%)** | **0.048** |
| Atopy biomarkers: |  |  |  |
| - Eosinophils, percent | **2 [1-3]** | **4 [2-7]** | **0.02** |
| - Eosinophils, cells/μL | **140 [50-240]** | **300 [190-580]** | **0.006** |
| - Total IgE, UI/mL | 501 [169-839] | 195 [80-704] | 0.59 |
| Subspecialist care: |  |  |  |
| - Pediatric Pulmonology | 20 (68.9%) | 84 (61.8%) | 0.47 |
| - Allergy / Immunology | 11 (37.9%) | 72 (52.9%) | 0.14 |
| Severe exacerbations: |  |  |  |
| - Any events<sup>2</sup> | 16 (55.2%) | 71 (52.2%) | 0.77 |
| - Events in the past year<sup>3</sup> | 1 [0.5-2.5] | 1.5 [1-2.5] | 0.31 |

Numbers represent mean (SD) or median [interquartile range] for continuous variables, and n (%) for categorical variables. ACT: Asthma control test (ages $\geq 12$ years, range 5-25). C-ACT: Childhood asthma control test (ages 4-11 years, range 0-27). <sup>1</sup>Defined as C-ACT or ACT $\leq 19$ . <sup>2</sup>Number of patients with $\geq 1$ asthma-related ED visit, hospitalization, or systemic steroid course from January 2018 to December 2019. <sup>3</sup>Rate among those who had at least one event.

